# Exploring the acceptability and feasibility of a blended imagery-based digital intervention for self-harm in young people (Imaginator 2.0): a co-produced qualitative study

**DOI:** 10.1101/2025.09.11.25335501

**Authors:** Lindsay H. Dewa, Athina Servi, Emily Gardner-Bougaard, Saida Jabril Mohamed, Aaron McDermott, C. Nusaybah, Ben Aveyard, Rachel Rodrigues, Adam Hampshire, Nejra Van Zalk, Martina Di Simplicio

**Affiliations:** School of Public Health, Imperial, London, UK; NIHR Imperial Biomedical Research Centre, London, UK; Department of Brain Sciences, Imperial, London, UK; Dyson School of Design Engineering, Imperial, London, UK; West London NHS Trust, London, UK; School of Psychology, University of Roehampton, London, UK

**Author notes:** Corresponding author: Dr Lindsay Dewa, School of Public Health, Imperial College London, London, UK 020 7594 0815.

**Keywords:** Digital mental health, self-harm, young people, Imaginator, co-production Word count: 5550

## Abstract

**Background:** Young people aged 12-25 experience self-harm, but interventions that are both acceptable and effective in reducing self-harm behaviour are rare. Digital interventions can be effective; however, they lack long-term engagement and are not embedded with the voice of young people. The Imaginator intervention is a novel, brief, blended intervention with a co-designed app that uses mental imagery to help young people manage self-harm. Our study aims to explore the acceptability and feasibility of the new Imaginator blended intervention (Imaginator 2.0) with young people.

**Methods:** We conducted a co-produced qualitative study with a purposive sample of young people who had used the Imaginator 2.0 intervention in West London NHS Trust. We conducted semi-structured interviews online using Microsoft Teams from April 2022 to December 2023. Three young people with lived experience of self-harm (i.e. co-researchers) informed every stage of the research cycle from design to dissemination. One co-researcher was trained in how to conduct semi-structured interviews with young people with mental health difficulties and subsequently conducted three interviews. Themes were constructed across three 2-hour virtual meetings with three co-researchers and the rest of the team. We presented the themes in a co-produced thematic map.

**Results:** Ten participants were interviewed (all female). There were four themes: Imaginator 2.0 therapy impact, mental imagery efficacy and limitations, app experience and engagement, and Imaginator 2.0 expectations and need for improvement. The app was found acceptable as a support tool around self-harm and a useful addition to therapy sessions. All app functionalities were mentioned as helpful by at least a few participants. Mental imagery was deemed a helpful aspect in reducing self-harm, although it was not always enough in moments of crisis. Overall, participants who experienced Imaginator 2.0 felt it had a strong impact on their emotions and behaviour and appreciated the therapist-patient relationship. Nevertheless, most participants felt that the app needed to be better integrated into the therapy and that app features should have more options for personalisation.

**Conclusion:** Overall, these findings support Imaginator as a brief, cost-effective early intervention that can be offered to young people when they first access services to help manage and contain self-harm. Our next step will be to conduct a randomised controlled trial (RCT) to compare Imaginator 2.0 to treatment as usual.

## Introduction

Reducing self-harm behaviour in young people is a worldwide priority. The lifetime self-harm prevalence is 20% in young people aged 16-25 years old (Lucena et al., 2022). Self-harm is defined as an intentional act of self-injury or poisoning that helps express distress (NICE, 2013). It is associated with several factors including poor health, cognition, and increased suicidal ideation, behaviour, and attempts (Mars et al., 2014; Ribeiro et al., 2016). Twenty per cent continue self-harming after 5 years, with increased severity and risk, with YP who self-harm are 17 times more likely to die by suicide (Morgan et al., 2017) The majority of those who self-harm do not access mental health services,(Geulayov et al., 2018) although in recent years, UK primary care self-harm presentations have exceeded expected rates. Therefore, there is a need for interventions that meet the high demand.

Psychosocial interventions have shown some success in reducing self-harm in young people. For example, cognitive behavioural therapy (CBT) and dialectic-behavioural therapy-based psychotherapy (DBT) have had low to medium effects on decreasing self-harm behaviour (Kothgassner et al., 2020), and can be expensive and lengthy. Whereas digital interventions for self-harm are scalable and could be crucial to help increase reach. However, only two digital interventions for self-harm have been evaluated for acceptability and feasibility: Blue Ice and ERITA. Blue Ice builds on CBT and DBT to help young people manage emotions and prevent self-harming behaviours (Grist et al., 2018; Stallard et al., 2016). It was co-designed with young people aged 12-17 and was found acceptable in two groups of young people (Cliffe et al., 2023; Grist et al., 2018). ERITA is a 12-session web-based emotion regulation programme targeting self-harm that also includes asynchronous contact with a dedicated clinician, which is cost-intensive. ERITA was also deemed acceptable. Despite these promising results, only ERITA has shown an advantage over standard care in reducing self-harm in adolescents (Bjureberg et al., 2023; Stallard et al., 2024). Overall, young people favour interventions supported by some form of human contact (Hollis et al., 2018). The lack of integration between the app and therapeutic work was also suggested to as a limitation of Blue Ice (Stallard et al., 2024). There is, therefore, a need for a co-designed digital intervention that is blended with face-to-face care, scalable, and cheap to deliver to give optimal acceptance, effectiveness, and cost-effectiveness to be successfully implemented into practice.

Imaginator is a blended digital intervention combining a co-designed smartphone app with three face-to-face therapy sessions and five follow-up phone calls (Di Simplicio et al., 2020)(Figure 1). It centers on Functional Imagery Training (FIT), which trains young people to use mental imagery to visualize alternative adaptive behaviours to self-harm. Young people visualise self-harm before engaging in the behaviour and that this drives the behaviour, which makes mental imagery a promising target for intervention (Ji et al., 2024; Lawrence et al., 2023; Witt et al., 2024). Imaginator has demonstrated early acceptability and efficacy in a small group of young people aged 16-25 (Di Simplicio et al., 2020). However, following clinicians and young people’s feedback, further co-design and development are needed to optimise Imaginator’s accessibility to younger adolescents, update its interface and features and make it personalisable to the user. Our study aim was to work in partnership with young co-researchers with lived experience to explore young people’s perspectives on the acceptability and feasibility of the newly co-designed Imaginator 2.0 intervention.

**Figure 1:**
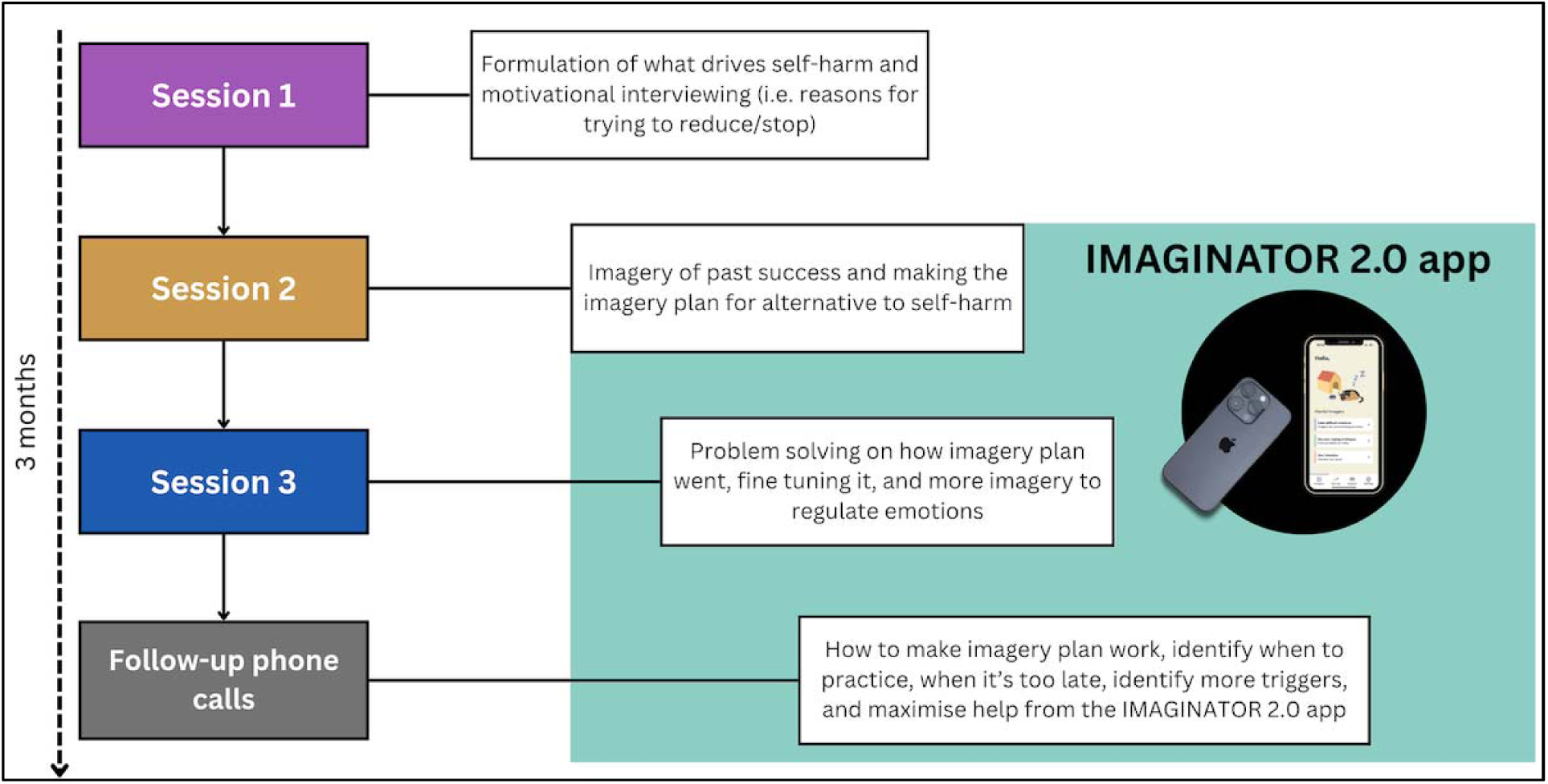
Imaginator overview

## Methods

### Design

This co-produced qualitative study was embedded within the wider co-design and early mixed methods evaluation of Imaginator 2.0 (Servi et al., 2025) (Figure 3). Participants completed a series of questionnaires at baseline and follow-up after receiving the Imaginator 2.0 blended digital intervention. A participatory lens was applied to the qualitative study, whereby power and responsibility were shared with a group of young people throughout the study. The consolidated criteria for reporting qualitative research (COREQ) (Tong et al., 2007) helped guide the reporting of this qualitative study (Staniszewska et al., 2017) (Appendix A).

### Co-production

We worked together in partnership with young people with experience of self-harm throughout all research stages (Ting, 2023) (Figure 3). The initial version of Imaginator was co-developed with a group of four young people who agreed that there was a need for a blended digital intervention that helped safely manage self-harm in young people. This initial version was piloted and reported (Di Simplicio et al., 2020). We needed to further update the intervention and brought new voices in (under 18) to co-design the newer Imaginator 2.0. We advertised for a new group of young people aged 14-25 to join the team on August 2021 via existing groups and four young people aged 17-24 applied as co-researchers. The co-researchers informed the funding application and after funding was secured to develop Imaginator 2.0, they planned and co-facilitated the app co-design workshops (reported separately) (Figure 2) and informed the feedback interview topic guide used as part of this qualitative study.

**Figure 2:**
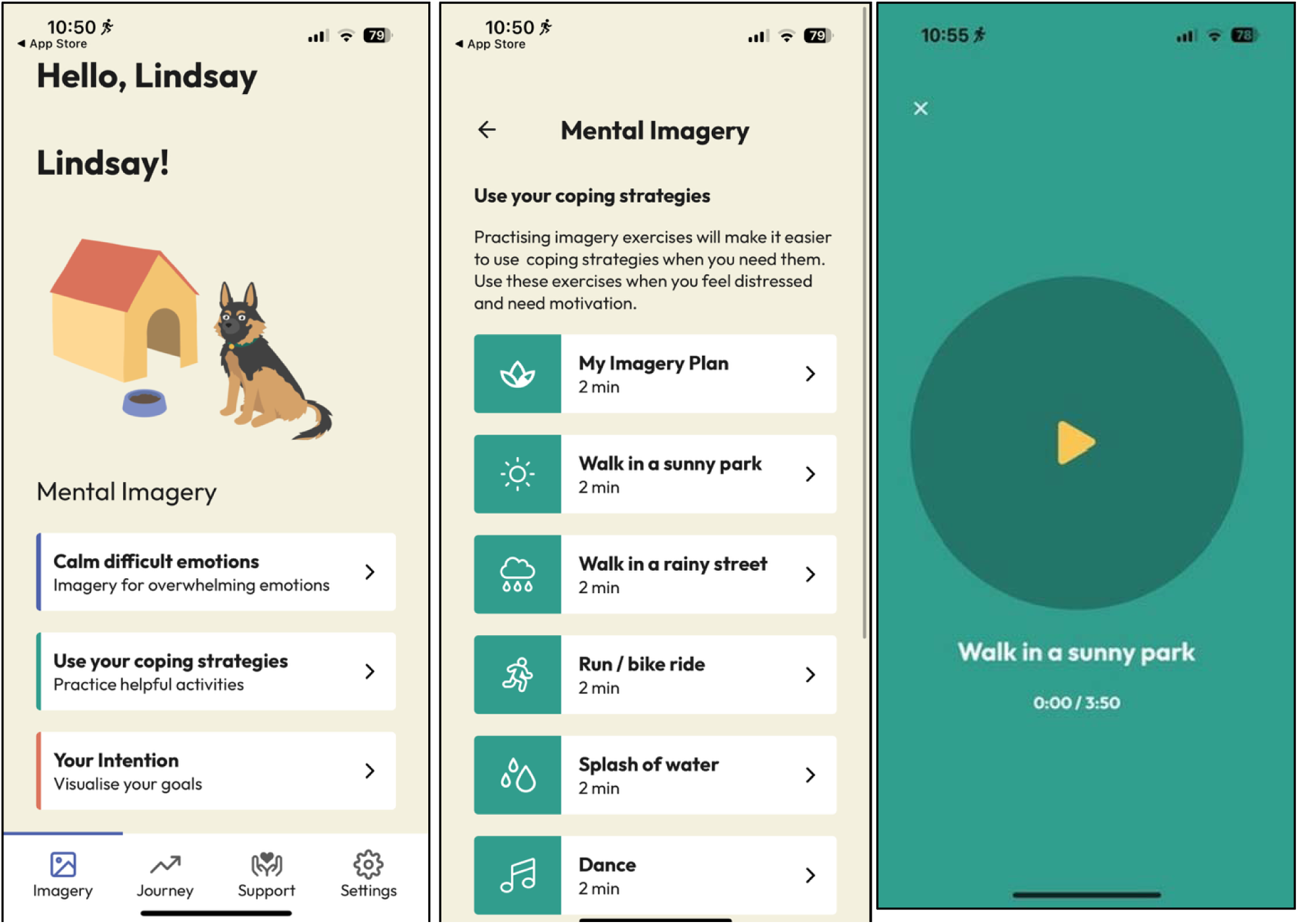
Imaginator 2.0 App screenshots

**Figure 3:**
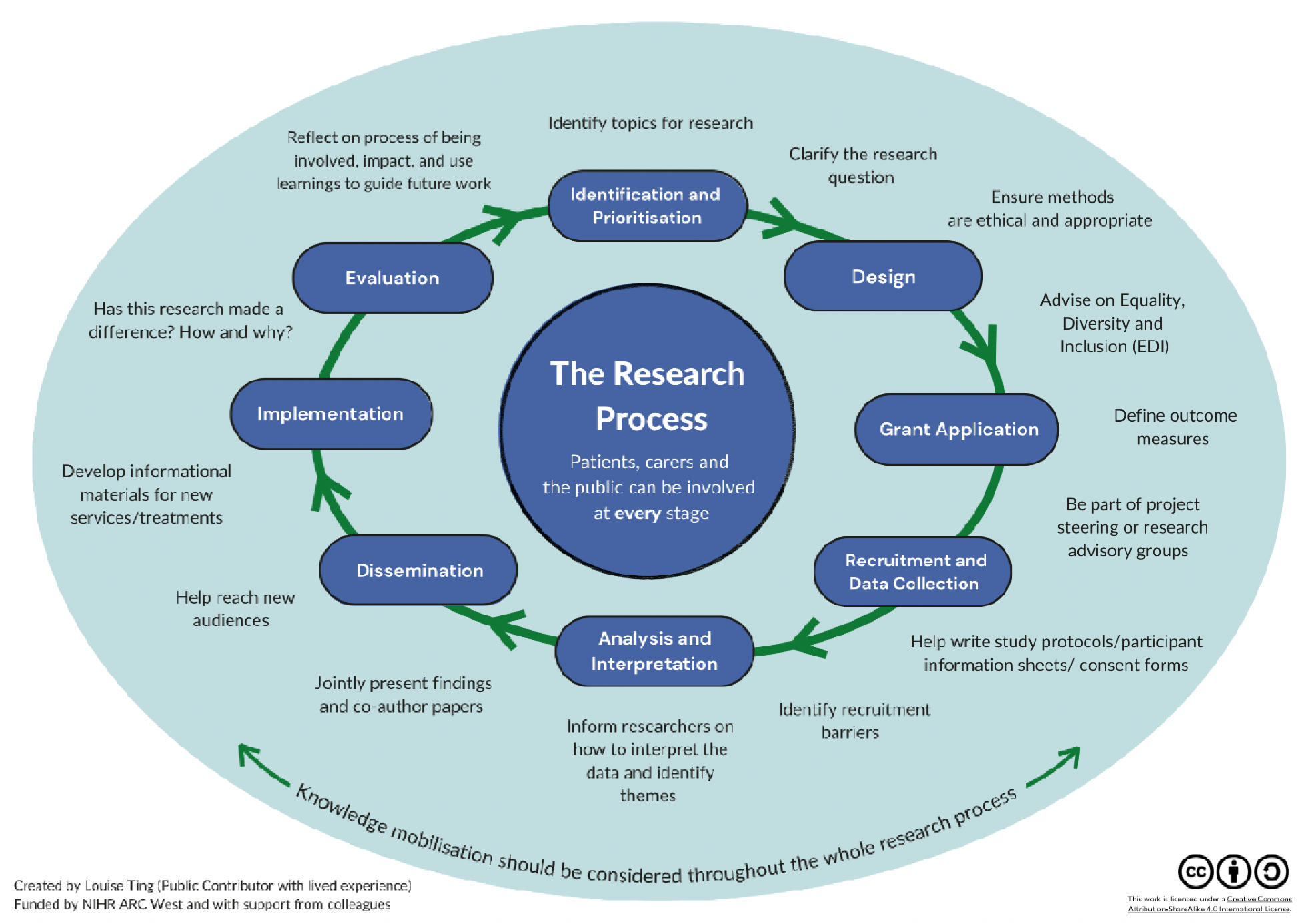
Research cycle

Three co-researchers were trained in conducting semi-structured interviews with young people with lived experience of mental health difficulties and self-harm, and basic thematic analysis through a pre-established training package (L. Dewa, C. Crandell, et al., 2021; L. Dewa et al., 2019; L. Dewa, A. Lawrence-Jones, et al., 2021). One co-researcher (SM) then conducted three interviews using an established three-pronged approach: 1) shadowing the researcher (AS) and debrief, 2) co-researcher interviewing with researcher (AS) shadowing and debrief, and 3) conducting the interview alone with researcher support. One co-researcher (SM) coded her own transcripts to help improve accuracy, nuance and quality of analysis. We had three 2-hour virtual meetings together as a team, co-creating the themes and thematic map. Finally, three co-researchers are co-authors of this paper. All co-researchers and co-designers were reimbursed with £25 an hour in accordance with NIHR guidance.

### Participants

Participants were recruited to take part in the Imaginator 2.0 intervention from adult Mental Health Integrated Network Teams (MINT) and child and adolescent mental health services (CAMHS) within West London NHS Trust if they were aged 12-25 years old, experienced at least 2 SH episodes lifetime or at least 5 SH episodes in the past year and currently reporting self-harm urges. Further details of the participants are explained elsewhere (Servi et al., 2025). All participants who completed the Imaginator 2.0 intervention for approximately 12 weeks and completed the follow-up assessments were approached to take part in the qualitative interviews. Written consent was obtained before the interviews. Participants were reimbursed with £10 per hour for participating in the interview.

### Data generation

Interviews were conducted online on Microsoft Teams by one researcher (AS) and one co-researcher (SM) (see PPI section) between April and December 2023. A topic guide was co-produced with the co-researchers and covered the following areas: experience of the therapy, the use of mental imagery, the Imaginator 2.0 app and integration of the therapy and app (Appendix B). All interviews were video and audio recorded, and transcribed verbatim by the researcher (AS). Interviews ended following the intended recruitment target rather than saturation. Interviews lasted between 45 and 60 minutes. Interview transcripts were imported into Imperial College London OneDrive ready for analysis. They were not returned to participants.

### Analysis

Inductive thematic analysis was conducted and guided by Braun and Clarke’s stages of thematic analysis, and the lead author’s established a co-produced analysis approach (L. Dewa, A. Lawrence-Jones, et al., 2021; L. H. Dewa et al., 2021; L. H. Dewa et al., 2019). One co-researcher (SM) and two researchers (AS, EG) familiarised themselves with the data by reading and re-reading the transcripts. The co-researcher (SM) conducted line-by-line coding across 3 transcripts, which was double-coded by a researcher (AS). The researcher (AS) then coded 7 transcripts, which were double-coded by another researcher (EG). All three (AS/EG/SM) then compared the codes to the initial coding framework and added their new codes. All codes were added to a Trello board. All researchers and co-researchers (AS, EG, LD, SM, MS, NC) met twice online using Microsoft Teams on February 28^th^ and 13^th^ March 2024 for a 1-hour workshop to review, refine and group the codes into themes. We split into pairs to group the codes into sub-themes by combining areas of similarity and difference under an initial sub-theme name.

Codes were then further refined, consolidated, and combined as part of a continuous process until we had sub-themes only and no repetition among the sub-themes. We then assigned sub-themes to a provisional theme name and transferred all sub-themes and themes into Miro to create a visual thematic map. We met a further time online on 3^rd^ April 2024 to further consolidate the theme and sub-theme names so that they made sense in isolation, to link the sub-themes and themes and to co-produce the final thematic map.

### Ethics statement

The authors assert that all procedures contributing to this work comply with the ethical standards of the relevant national and institutional committees on human experimentation and with the Helsinki Declaration of 1975, as revised in 2008. All procedures involving human subjects/patients were approved by West of Scotland Research Ethics Committee (22/WS/0087).

## Results

### Participants

Ten participants completed the qualitative interviews (N = 10/27) of those who started therapy). All were female (100%). Participants varied in age (*mean* = 20 years old, *sd* = 4.14), ethnicity (70% White), and sexual orientation (30% Heterosexual) (Table 1; Appendix C). Just under two-thirds (60%) were still in education, while some (30%) were employed. Seven (70%) had suicidal thoughts, and nine had self-harmed in the past month (90%). All completed therapy as per protocol.

### Themes

There were four main themes: Imaginator 2.0 therapy impact, mental imagery efficacy and limitations, app experience and engagement, and Imaginator 2.0 need for improvement (Figure 4).

**Figure 4:**
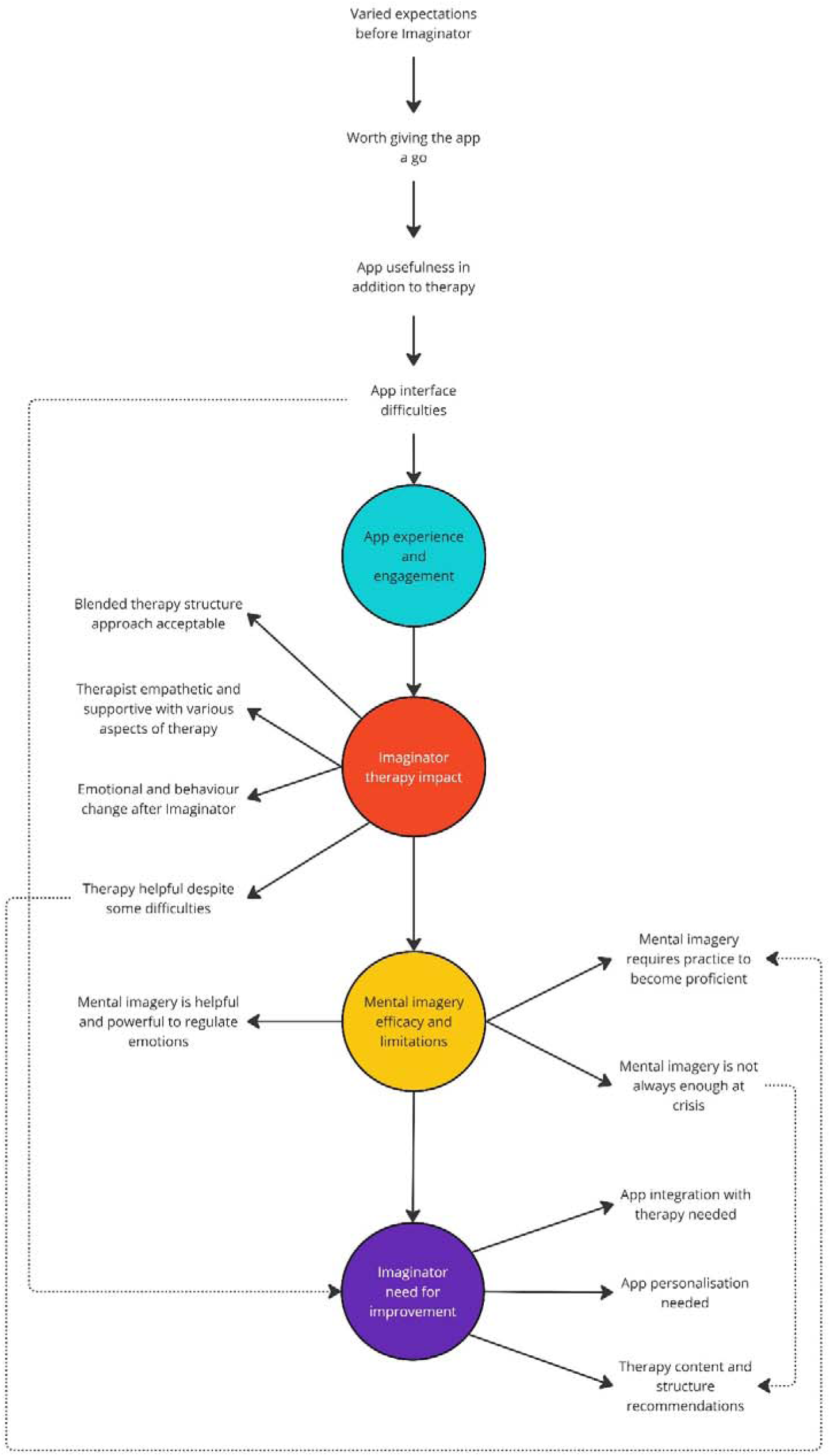
Co-produced thematic map

#### Imaginator 2.0 therapy impact

The impact of the Imaginator 2.0 therapy was twofold: 1) the impact the therapy had on the participants’ emotions and behaviour, and 2) the impact of the therapist-patient relationship on participants’ emotions and understanding of self-harm. Most participants felt having the Imaginator 2.0 therapy sessions with the additional support of the app made them feel better following the therapy sessions. For example, some participants reported fewer mood fluctuations, recognising emotions better, improved coping skills, and less self-harm behaviour after therapy. One participant reported that life had improved following the therapy sessions and that the therapy helped validate their feelings. There were also reported unexpected positive consequences of having the therapy for some, including having a better relationship with food, improved relationships, personal growth, and developing the realisation that cannabis was bad for their health.

> *“I’m kind of recognising when I’m thinking negatively. Before, it was kind of just my natural state and that’s what you believe because you believe what your brain tells you, whereas now I can kind of change it to a positive image or positive framework. I can still notice that I’m thinking negatively and that it’s not necessarily real. And I think that’s probably the biggest change. I guess I feel proud about it, and also it makes me feel like I’ve got a solid ground to try and change things now.”*

*Participant 2*

> *“One of the goals I set was to always ask for help. Asking for help now is a lot easier than before. Now I know how to ask for it. Before the therapy, I was waiting for someone to ask me if I were ok. Now I can just go up to people if I’m upset and just say this happened, and they’d be grateful that I told them first.”*

*Participant 3*

> *“I actually had the urge to put a blade to my wrist, but I started thinking about the stuff that I learned during these sessions and was able to redirect my chain of thought. I was actually pretty pleased with myself because I haven’t been able to redirect my chain of thought before.”*

*Participant 6*

All participants loved having someone to talk to, and the strong social connection with the therapist was one of the most helpful aspects of the therapy. For example, participants reported varied aspects of therapeutic alliance and having a strong social connection, including that the therapist was helpful, understanding, non-judgmental, empathetic, made them feel heard, and gave them space to explore things they wanted to discuss. Similarly, some reported that the therapist was not only supportive, but they delivered the therapy in a structured manner that was personalised. For example, one participant reported being able to brainstorm alternative coping strategies to self-harm, and that the therapist was adaptive to their needs.

> “*When I started the therapy, I knew I needed help. Every time I would be doing therapy, my therapist always reminded me of the amount of growth that I’ve accomplished. And I felt that it really did mean a lot to me because sometimes you don’t even realize how much growth you have accomplished. I had a lot of emotional growth. So, I think that it has helped me a lot. There were many times where I would just look forward to [therapy sessions] when I was having a bad week. I am really happy I took this course, and I would always tell my therapist how much I appreciate it because it really helped me”*

*Participant 1*

> *“I think it was an interesting experience because I was very honest about SH in a way that I hadn’t ever previously had the option to be. And I think that was really powerful. […] SH has always been the elephant in the room. Everyone did not want me to mention it or talk about it. […] So, it was really important for me to have that opportunity to explore not just in the session itself, but in the build-up to the session as well and to really interrogate it […] Having the space to talk about SH. A non-judgmental place. Not seeing people recoil in horror.”*

*Participant 7*

The therapy structure (3 1-hr in-person sessions and 5 phone calls) was mostly acceptable to most participants. However, participants had mixed views on the delivery mode of the therapy, as some liked the blended approach, but others preferred more in-person therapy sessions.

> *“I have actually liked the format because it gives that kind of opportunity where if some people aren’t comfortable talking in person, they have the opportunity to do it online because I feel like most therapies expect that person to be in the appointment in person all the time. But I feel like when it’s when some appointments are online, it kind of more considerate because they might be like having a bad time talking about why they’re in…therapy and then they might just need to break at home and the online sessions do that.”*

*Participant 10*

A few participants reported negative impacts of the therapy. For example, some lost motivation for the therapy sessions, it could be emotionally draining and feeling worse after the final therapy session, knowing it was the last one.

### Mental imagery efficacy and limitations

Overall, Imaginator 2.0 was deemed acceptable as a blended intervention to help reduce self-harm. Most participants reported that imagining alternative things to self-harm was a powerful way to help regulate emotions. Mental imagery content ranged from reminiscing feelings of past successes to scenarios about the future that felt joyful. These mental images led to positive emotions such as feelings of joy and happiness, motivation and a sense of calmness. Participants reported that regulating emotions early through mental imagery was helpful to prevent getting overwhelmed, ruminating and self-harming or engaging in other unhelpful behaviours.

> *“If you had asked me before the therapy, I’d have told you that this is rubbish and would not work. I did absolutely go into it with an open mind because there was no point in doing it otherwise, but I really actually found it so useful. I’ve done so much CBT imagery where you picture yourself on a boat etc. That was stressful. Guided meditation does not work for me at all. I mean at some points almost I was forced to come up with these images which was really kind of a powerful technique for me, and it was so different to anything I’d ever experienced or considered experiencing and I really enjoyed it.”*

*Participant 7*

> “*There was a time where I struggled to get out of bed sometimes, so we thought of an image that I could use to get out of bed. So, I think it was quite nice trying to figure out what could work. I think this part of the therapy sort of reminded me of my nieces and nephews (that was my image) and that is why I liked it. It made me happy, and it was just nice to be reminded.”*

*Participant 8*

> *“It’s helped me stop doing a lot of bad things I could have done. There’s one situation it helped because I really wanted to get drunk, like I can’t walk drunk. And then that mental imagery came up, my partner helped me get that mental imagery running in my mind, and then I just started to imagine what would have happened if my family saw me for example, hospitalised from getting alcohol poisoning or something like that. It was a weird thought, and it just helped me realize just maybe let’s not do this type of thing.”*

*Participant 10*

However, some felt that mental imagery was not always enough when they were already at crisis point because they were too overwhelmed. Two participants explained that despite this, the exposure to the mental imagery helped to distract during the moment of crisis. Furthermore, despite the positive response to mental imagery, some participants still found mental imagery difficult, tiring and that it required a lot of effort. However, most felt that mental imagery got easier the more they practiced.

> “*I’m trying to use visual imagery, but I struggled with that. I struggled with trying to take my mind there because my mind’s quite busy naturally, but then we worked on using prompts for things around my space to help me get to a space that I find like safe for a good distraction. And that was better.”*

*Participant 5*

> “*I don’t think it ever made me feel better. It helped me in the moment. I knew that my problems weren’t fixed by doing it, but I knew that I could get through the next hour or so and not be stupid or overwhelmed. It helped in terms of self-harm because when I wanted to hurt myself, I used it instead of hurting myself. Whether it works depends on how quick I am to use the app because if I’m a bit late and I’m already overthinking its unlikely to work but if I catch it when I’ve only started to think about my thoughts getting a bit loud it’s more likely to work. It never made me feel worse it just made me annoyed sometimes.”*

*Participant 9*

### App experience and engagement

Most participants reported they wanted to use the Imaginator 2.0 app in the first instance because they thought it looked useful and appealing. For example, some participants felt the app was appealing because of the calming colours, the dog mascot that followed them through the app, and because it was simple to use. Most participants reported that the Imaginator 2.0 app’s core features (the imagery plan and imagery audios), as well as the crisis support feature, were helpful. The imagery plan and audios helped them identify, monitor, and manage their emotions, helped with sleep, helped to stop rumination and were described as relaxing.

However, some did not like some of the audio voices (e.g., “American accent is not relaxing”). Most participants liked the immediate support feature that was offered to support in crisis or following self-harm (e.g., Shout – 24/7 crisis text line).

> *“I think I used the audios whenever I felt down, I think that was almost everyday of the week.”*

*Participant 3*

> *“Anytime I’d feel like a really strong urge to throw up, in the back of my mind, I would just pull out the rainy audio and that would help calm me and calm my mind.”*

*Participant 4*

> *“I found [the audios] quite helpful to be honest. I find them quite calming, especially when I’m overwhelmed or triggered or anything. I use them mostly at night because that’s when I’m usually left alone with my own thoughts, which is not always nice.”*

*Participant 6*

> *“One night I was kind of in that spiral. I started to think about hurting myself and I was like, that sounds amazing right now to just hurt myself and I wanted to, and I felt my mood decreasing. […] And then I was just on my phone. I was like, “you know what? Let me just go on that. Let me just listen to that coping mechanism audio” and then I listened to it and somehow it helped. It just made me realize I don’t need to hurt myself.”*

*Participant 10*

Participants had mixed views on the remaining features (the mood tracker, journal and goals tracker). For example, most participants loved the mood tracker because it was helpful to understand their feelings and emotions, whereas a few did not like tracking their mood and did not find it helpful. Most participants did not like the journal, and goals tracker features, resulting in some participants not using them. Additionally, most participants reported not liking at least one aspect of the app such as the interface, content, or usability. A few participants also reported some interface difficulties to using the Imaginator 2.0 app including being “buggy” and “clunky” and not receiving notifications. One person reported that they did not like the Imaginator 2.0 app enough to download it again after they changed phones.

> *“Where it was a bit annoying was when my phone turned off, I couldn’t listen to [audio] because then if I fall asleep, my phone’s going to be on all night and not charged. But listening to it a couple of times just before I went to bed was really helpful because it kind of quietened down the anxious thoughts.”*

*Participant 2*

> *“I had to tap the screen of my phone when listening to the audio to keep the phone open. If it closed the voice would stop. I don’t want to sit there and keep tapping on my phone to make sure the screen is on.”*

*Participant 8*

#### Imaginator 2.0 need for improvement

Participants reported varied expectations of Imaginator 2.0 (therapy and app) before they started the intervention. Some had an open mind, whereas some had low expectations of the app and did not feel it would help them. Despite this expectation, most reported that Imaginator 2.0 did help them to regulate their emotions (amongst other things). However, all participants still felt there was a need to improve Imaginator 2.0 (therapy and app). With the therapy, some participants reported still having difficulty controlling emotions and thoughts, and therefore would like more time dedicated to supporting emotion regulation through mental imagery within the intervention. Similarly, some participants reported more time to explore the root cause of self-harm would have been helpful, and to have more 1-1 sessions face-to-face.

> *“The therapists contact number could be there in the future e.g. if you were doing the mood tracking you should have the option to share with your therapist”*

*Participant 5*

> “*What would have been good to have is a face-to-face session in the end after the phone calls or a variation like that. I think that having another in person session would have been helpful because there is do much body language that goes on.”*

*Participant 7*

> “*Instead of making someone think on the spot about a memory [reference to the “imagery of a past success” exercise in Session 2], because this usually doesn’t work, maybe set it as something they have to do at home, maybe with a childhood toy, or maybe a certain music. And they kind of have to think about that and then kind of going back in.”*

*Participant 10*

Some participants reported that the Imaginator 2.0 app and the therapy sessions felt like two separate entities that were operating in isolation. For example, some indicated that they did not have enough exposure to the app, that they did not use the app during the sessions or discuss their app usage before the therapy within the sessions. Of these participants who reported this, all felt there was a need to better integrate the Imaginator 2.0 app with the therapy.

> *“I wanted to use [the app] but I didn’t. Maybe because of a lack of motivation. Nothing to do with the app. I was introduced to the app and my therapist told me to use it in my own time. I feel that if it was implemented in the therapy, I would have used it more. I would always forget.”*

*Participant 1*

> *“I think it was a really helpful app, but I think for me it felt quite disjointed. There was the therapy and then there was the app. And so, I wasn’t really sure where to bring the app in because a lot of my little tasks or homework tasks wouldn’t really relate to the app.”*

*Participant 2*

Participants proposed multiple and varied changes for the Imaginator 2.0 app to make it better for young people in the future. These changes varied between app content, structure and appearance but centred on the need for better personalisation. For example, some participants reported they were not comfortable hearing their own voices on the app, and therefore one participant suggested having someone else record the imagery plan. Others suggested having a wide range of voices (e.g., instead of using the American voice), and having longer audios.

Reported app appearance changes included the option to choose a mascot (e.g., not just a dog), varied and tailored helplines and crisis support based on age and location (e.g., putting in postcode); more interactivity and having the therapy booklet integrated within the app.

> *“Maybe add something that can show your progress a bit better. Something more encouraging maybe or what happens when you complete your goals. Maybe it could integrate something about how there’s a reward, or that you can make your own reward for when you complete your goals, and then that goes at the end of the scale and it’s kind of more motivating to complete it.”*

*Participant 2*

> *“I am a dog person but maybe we could have a choice of which mascot we would like. […] I think the app is overall impersonal. I would also like to choose the colors on my phone. I read better with a darker background and white letters (like the dark mode). Reading on a white background gives me migraines. So, if there was an option to change that, it would be helpful.”*

*Participant 8*

## Discussion

This study aimed to explore the acceptability and feasibility of a new blended mental imagery-based digital intervention for self-harm in young people called Imaginator 2.0. There were four themes: Imaginator 2.0 therapy impact, mental imagery efficacy, app experience and engagement, and limitations and Imaginator 2.0 need for improvement. Overall, participants felt that Imaginator 2.0 was acceptable and feasible and it importantly had a positive impact on regulating emotions and subsequently self-harm, in line with other self-harm apps (Cliffe et al., 2023; Grist et al., 2018),. Of note was the strong therapeutic alliance with the therapist, and how the core app features (e.g., imagery plan and audios) helped identify, monitor, and manage their emotions and behaviour. However, several technological concerns would need to be addressed to make the intervention successful in the future. These included interface difficulties, and mental imagery-based techniques not being easy to execute, or not being enough at the point of crisis, as participants would either forget or find mental imagery too hard to use. However, despite these issues, there is potential for Imaginator 2.0 to be used in practice to regulate emotions and reduce self-harm, particularly before becoming too overwhelmed to fully benefit from the intervention. Recommended changes included having more time dedicated to practice mental imagery and emotion regulation within the app, more personalisation and better integration of the therapy with the app.

Participants’ engagement with the Imaginator 2.0 app appeared to be influenced by the app’s design, features, and personalisation. This is consistent with previous findings on digital mental health interventions, suggesting that the most effective apps are those that can be tailored to individual users, aligning with their personal care goals and needs (Torous et al., 2020).

Participants’ concerns around sustained use and motivation echoed broader challenges often encountered in digital mental health interventions, particularly among young people, where dropout rates remain high and daily use is difficult to maintain without strong therapeutic scaffolding (Hollis et al., 2017). As participants reported, sometimes the app appeared to be separate from the therapy sessions, impacting their engagement with it. In contrast, participants found therapy sessions to be validating and transformative, aligning with evidence that therapeutic alliance and co-construction of meaning can be key mechanisms of change in psychological interventions for self-harm (Dunster-Page et al., 2017; Huggett et al., 2022).

Having the space to talk and explore their self-harm was a new experience for most participants, as self-harm is often stigmatised and overlooked. Participants’ feedback highlighted that their therapist in the Imaginator 2.0 intervention focused on building trust, which was integral to forming an alliance. This underscores the importance of creating a safe and reassuring environment, particularly for individuals who may feel apprehensive about opening up about their emotions and feelings (Sass et al., 2022).

Within the overall Imaginator 2.0 intervention, mental imagery was identified as a particularly helpful aspect, offering participants a creative and emotionally resonant way to process thoughts and feelings related to self-harm. Similar findings have been reported in qualitative research on imagery-based treatments for anxiety in bipolar disorder. For example, participants expressed broadly positive experiences with the intervention and described improvements in anxiety management, agency, and daily functioning following the use of mental imagery techniques (Elkington et al., 2024). Previous research also highlights the unique power of mental imagery to evoke stronger emotional responses than verbal processing alone (Blackwell et al., 2025) and may therefore be especially effective in reducing self-harm urges. FIT focuses on imagining a coping strategy plan and strengthening the motivation to put this in action (May et al., 2015). Interestingly, several participants in our study reported that using mental imagery supported them directly to regulate emotions. However, they also expressed a need for more support around emotion regulation strategies within the therapy sessions. Future studies may dissect whether Imaginator 2.0 works more via motivational or emotional regulation mechanisms and what components are needed to maximise efficacy. Moreover, some participants described that while targeting self-harm urges mental imagery also helped reduce other behaviours such as disordered eating, alcohol use, and cannabis consumption. Mental imagery has been shown to activate similar neural and emotional systems as real-life experiences (Ji et al., 2019; Pearson et al., 2015), making it a powerful tool for influencing affective states. Its ability to simulate positive outcomes or coping strategies may enable individuals to reframe distressing experiences, reduce impulsive behaviours, and enhance self-control across a range of maladaptive behaviours (Andrade et al., 2016; May et al., 2015).

Previous research has found that guided imagery can improve regulation of craving and emotional distress in substance use disorders (Bakou et al., 2021; Kober et al., 2010; May et al., 2015), supporting the notion that imagery-based techniques tap into transdiagnostic mechanisms. These findings underscore the potential for mental imagery interventions to extend beyond self-harm and support wider emotional and behavioural regulation.

Participants acknowledged the preventative value of mental imagery in helping them reduce the likelihood of reaching a crisis-point, however, they also emphasised that it was not always enough during moments of crisis. One possible explanation is that mental imagery relies on a baseline level of cognitive and emotional regulation to be effectively accessed and implemented (Skottnik & Linden, 2019). Individuals with greater mental imagery abilities, such as vividness and controllability, tend to have higher levels of dispositional optimism and well-being (Blackwell et al., 2013; Odou & Vella-Brodrick, 2013), which may reflect their capacity to engage in adaptive emotional processing. However, during episodes of intense distress, cognitive resources are often compromised, and emotion regulation becomes more difficult, making it harder for individuals to generate or maintain positive mental images (Andries et al., 2024; Hoppe et al., 2022). This highlights the need for additional, easily accessible crisis strategies to complement imagery-based tools, ensuring support is available across varying emotional states. Furthermore, participants reported the need for intervention improvements, such as increased personalisation, extra face-to-face sessions to support content delivery, and better integration of the app with therapy sessions. This supports the growing view that self-harm is a highly heterogeneous behaviour and there is a recognition that digital interventions need to be extremely personalised (Moran et al., 2024) and should be meaningfully embedded within a wider therapeutic framework (Nelson et al., 2024). There is therefore a need for further development and testing of Imaginator 2.0 with a RCT to compare it to treatment as usual.

Future research may include delivering Imaginator 2.0 outside mental health services due to its brevity and delivery by low-intensity practitioners, and its impact could be extended to other dysregulated behaviours (e.g., anger, impulsivity).

### Strengths and limitations

This qualitative study enabled a nuanced exploration of participants’ experiences with our Imaginator 2.0 intervention. Our reflexive thematic analysis approach ensured that the findings were grounded in participants’ voices and contributed to a growing but still limited body of literature on novel, blended interventions for self-harm. Most importantly, young people with lived experience were involved in all stages of research, including data collection, analysis, and dissemination (Ting, 2023). Even though small, the sample of this study was diverse, representative of the highly diverse urban West London area. Nevertheless, participants who took part in the interviews were typically those who completed most aspects of the intervention, resulting in a lack of insight into the experiences of individuals who disengaged early. This introduces a potential selection bias, as the sample may overrepresent those who found the intervention acceptable or beneficial.

## Conclusion

Overall, these findings support Imaginator 2.0 as a brief, cost-effective early intervention that can be offered to young people when they first access services, including during the waiting period for other therapeutic support, to help manage and contain self-harm. The app was found almost unanimously acceptable as a support tool for self-harm and a useful addition to therapy sessions. Participants’ experience showed idiosyncratic use and preferences for the different app functionalities and visual/engagement aspects. All functionalities were mentioned as helpful by at least a few participants (e.g. mood tracker, journal, goals tracker, different audio types), suggesting that they should stay in the final version of the app and that we managed to capture the diversity of needs of the population. While the app was well introduced by the therapists, most participants felt that it needed to be better integrated into the therapy, for example, to facilitate using it more often. Going forward, we expect to conduct an RCT to compare the effectiveness of Imaginator 2.0 treatment as usual on reducing self-harm in young people.

## Data Availability

Data is available upon request.

### Appendix A: COREQ

| Item No | Guide Questions/Description | Reported on Page # |
| --- | --- | --- |
| **Domain 1: Research team and reflexivity** |  |  |
| **Personal Characteristics** |  |  |
| 1. Interviewer/facilitator | Which author/s conducted the interview or focus group? | 8 |
| 2. Credentials | What were the researcher's credentials? E.g., PhD, MD | 1 |
| 3. Occupation | What was their occupation at the time of the study? | N/A |
| 4. Gender | Was the researcher male or female? | N/A |
| 5. Experience and training | What experience or training did the researcher have? | 8 |
| **Relationship with participants** |  |  |
| 6. Relationship established | Was a relationship established prior to study commencement? | 5 (reported in Servi et al., 2025) |
| 7. Participant knowledge of the interviewer | What did the participants know about the researcher? e.g. personal goals, reasons for doing the research? | 5 |
| 8. Interviewer characteristics | What characteristics were reported about the interviewer/facilitator? e.g. Bias, assumptions, reasons and interests in the research topic | 5 |
| **Domain 2: study design** |  |  |
| **Theoretical framework** |  |  |
| 9. Methodological orientation and Theory | What methodological orientation was stated to underpin the study? e.g. grounded theory, discourse analysis, ethnography, phenomenology, content analysis | 5 |
| **Participant selection** |  |  |
| 10. Sampling | How were participants selected? e.g., purposive, convenience, consecutive, snowball | 7 |
| 11. Method of approach | How were participants approached? e.g., face-to-face, telephone, mail, email | 7 |
| 12. Sample size | How many participants were in the study? | 9 |
| 13. Non-participation Setting | How many people refused to participate or dropped out? Reasons? | 9 |
| 14. Setting of data collection | Where was the data collected? e.g., home, clinic, workplace | 7 |
| 15. Presence of nonparticipants | Was anyone else present besides the participants and researchers? | N/A |
| 16. Description of sample | What are the important characteristics of the sample? e.g. demographic data, date | 9 & 36 (Appendix D) |
| **Data collection** |  |  |
| 17. Interview guide | Were questions, prompts, and guides provided by the authors? Was it pilot tested? | 30 (Appendix C) |
| 18. Repeat interviews | Were repeat interviews carried out? If yes, how many? | N/A |
| 19. Audio/visual recording | Did the research use audio or visual recording to collect the data? | 8 |
| 20. Field notes | Were field notes made during and/or after the interview or focus group? | N/A |
| 21. Duration | What was the duration of the interviews or focus group? | 8 |
| 22. Data saturation | Was data saturation discussed? | 8 |
| 23. Transcripts returned | Were transcripts returned to participants for comment and/or correction? | 8 |
| **Domain 3: analysis and findings** |  |  |
| **Data analysis** |  |  |
| 24. Number of data coders | How many data coders coded the data? | 8 |
| 25. Description of the coding tree | Did the authors provide a description of the coding tree? | N/A |
| 26. Derivation of themes | Were themes identified in advance or derived from the data? | 8 |
| 27. Software | What software, if applicable, was used to manage the data? | 8 |
| 28. Participant checking | Did participants provide feedback on the findings? | N/A |
| **Reporting** |  |  |
| 29. Quotations presented | Were participant quotations presented to illustrate the themes/findings? Was each quotation identified? e.g., participant number | 11-17 |
| 30. Data and findings consistent | Was there consistency between the data presented and the findings? | 18 |
| 31. Clarity of major themes | Were major themes clearly presented in the findings? | 9 |
| 32. Clarity of minor themes | Is there a description of diverse cases or a discussion of minor themes? | 9-18 |

### Appendix B: Feedback Interview – Topic Guide

#### Introduction to the interview

Thank you very much for meeting with me today. My name is [insert name] and I work at Imperial College London/as a co-researcher on this study. As you know we are carrying out a research study to explore how young people have found the Imaginator therapy that you’ve recently completed. We would love to hear about your experience with the therapy and the app, which is why we’re doing this interview. The interview is very informal and confidential. The only exception to this is if, after the interview, we feel your or someone else’s health or safety is at immediate risk because of something you have told us. If this happens we will inform a member of the mental health team immediately. We’ll always let you know first before we do this. Only myself and the direct members of my team will see the interview or hear it. Your name will not appear on anything that we write, and you will be notified before we publish the results. With your permission I would like to record the interview. This is so I can concentrate on your responses rather than being distracted by taking notes. All recordings will be destroyed once the project is complete. This should be seen as a safe space where you can talk openly about your experiences. You can stop and take a break at any point, and we can skip any questions you don’t feel comfortable answering. Have you got any questions before we start? Thank you we’ll make a start.

1. Therapy

- **Opening question:** First I want to ask you about your experiences with the therapy. This includes the 3 face-to-face sessions and the 5 telephone follow-up calls you had with your therapist.
- General feedback:

- *Can you tell me about a time where the therapy really worked—or didn’t work—for you? What happened?*
- *Can you tell me about something that you did in the sessions (in person or over the phone) that you particularly liked or disliked? Why was that?*
- **Set-up:** What did you think about how the therapy was structured? This might include for example, the length of the therapy (3 sessions + 5 follow-up calls) or the format e.g. whether you did it online or in-person.

- *How did this impact or not impact your experience of the therapy?*
- Impact of therapy on mental health:

- *When you think back to how you felt before you started the therapy and how you feel now … is there anything that changed?*
- *Can you tell me about a recent situation where you think you reacted differently because of the therapy? (How / why / what was different)?*
- *More broadly, is there anything you find yourself doing more / less of now? And how do you feel about this change? This might include self-harm or whether you drink alcohol or use other substances, your mood, thoughts and feelings around suicide, or anything else.* (prompt – how did the therapy help/hinder progress?)
- Therapeutic alliance:

- *Can you tell me about something the therapist said that stayed with you? That you particularly liked or disliked?*
- *Did you feel like you were looking forward to seeing them or receiving their call at any point? When was that / why?*
- *And what about the opposite (not looking forward to seeing them)?*
- *Do you remember a time when seeing the therapist made you feel in a certain way, good or bad? What happened?* (prompt – did you feel a sense of connection with the therapist, if so – why? If not – why not?)
- **Goal setting:** The therapy aimed to help you achieve specific goals that you set for yourself. Can you remember one of the goals you set? Can you tell me about an example situation when you felt the therapy supported you in reaching / not reaching the goal?
- **Using other coping strategies:** Another part of the therapy aimed to help you use different coping strategies other than self-harm. Can you remember an example when you used a different coping strategy, that you had discussed in the therapy? A time when trying a new coping strategy was useful or not useful?
- **Therapy ending:** How have you been finding managing without support from your therapist, now that the therapy has ended?
- **Any improvements:** Is there anything you feel could be improved?
2. Mental imagery

- **Opening question:** The therapy used a particular technique involving mental imagery where you imagined achieving your goals and using your coping strategies. What did you think of the mental imagery technique?

- *Can you give me an example of when you used (or still use) mental imagery? How did you find using it?*
- *What about a time when you tried but didn’t find it helpful or maybe it was difficult?* (prompts – what did you find useful or not useful about using the mental imagery techniques? Was it easy or difficult to use – and why do you feel this was? Did you use it as much as you intended to or not, and why was this? Try to get more than one imagery example)
- **App audios:** Did you use any of the mental imagery audios in the app?

- **If no:** Can you give me an example of how you practice / have practiced mental imagery without the app outside the therapy sessions (if you do)?
- **If yes:** *What did you think of the mental imagery audios in the app?* (prompts – What was the variety of audios like for you? Did you have any preferences on the audios? Why did you like them? Which ones didn’t you like, and why? Anything you missed in the audios? Would you add or change anything e.g. duration, voices, content?)
- **‘Use your coping strategies’ audios:** Open the app and the show the interviewee these mental imagery audios.

▪ *Could you give me an example of using these audios (if you did)? How did you find it?* (prompts - How were the length of the audios, did you use any types of imagery more or less, and why, or why not did you use them in this way?)
▪ *During the therapy did you create and upload an audio called ‘my imagery plan’ with your therapist (show on app)? What was your experience of doing this with your therapist? What was your experience of using this audio in the app?* (prompts – Was there anything you liked or disliked about this? Was this useful or not?)
- **‘Calm difficult emotions’ audios:** Show the interviewee these mental imagery audios in the app.

▪ *Could you give me an example of using these audios (if you did)? How did you find it?* (prompts - How were the length of the audios, did you use any types of imagery more or less, and why, or why not did you use them in this way?)
- **‘Your Intention’ audio:** Show the interviewee this mental imagery audios in the app.

▪ *Could you give me an example of using this audio (if you did)?* (prompts - How was the length of the audio, did you find it useful or not useful?)
- **Selecting the audios:** *Can you describe your experience around choosing which mental imagery audios to listen to?* (prompts – Were there any reasons you chose to listen to specific audios or not? If yes - what were these reasons?)
- **Categorisation of audios:** *How did you find the grouping and descriptions of the audios?* (prompts – was the scope of each category clear or not?)

▪ **If it wasn’t clear for them:** *How would you group and describe them?*
- **Impact of mental imagery:** Can you tell me about a time when using mental imagery made you feel better in terms your mental health, if that happened? This might include self-harm or whether you drink alcohol or use other substances, your mood, thoughts and feelings around suicide, or anything else. What happened and why do you think that worked or didn’t work? And what about any time it made you feel worse?
- **End of mental imagery:** Did you stop using the mental imagery once the therapy ended? If so – why did you do this? If not – why did you carry on using it?
- **Therapy and mental imagery:** *Do you have any feedback on how mental imagery techniques can be enhanced with the therapy?* (prompts – for example how can the therapy help to make mental imagery stronger/more vivid? How can the therapy help people to use the mental imagery techniques more?)
- **App and mental imagery:** *Do you have any feedback on how mental imagery techniques can be enhanced with the app?* (prompts – for example how can the app help to make mental imagery stronger/more vivid? How can the app help people to use the mental imagery techniques more?)
3. App

- **Opening question:** It’s been great talking to you about your experiences so far, thank you for sharing. We’ll move onto talking a bit about the app. Can you tell me of times you have been using the app? when was that and what did you do? How did you find it?
- **Exploring answers from User Experience Questionnaire (UEQ):** Only explore this if they scored low on any subscale of the UEQ. *In one of the questionnaires asking you about the app, you indicated that…:*

- If scored low on <u>attractiveness</u> subscale: *the app wasn’t very attractive*
- If scored low on <u>perspicuity</u> subscale: *the app wasn’t very easy to use*
- If scored low on <u>efficiency</u> subscale: *the app wasn’t very efficient*
- If scored low on <u>dependability</u> subscale: *the app wasn’t very dependable* (prompts - For example this could relate to how predictable the app was, whether you felt it was secure, or whether it met your expectations.)
- If scored low on <u>stimulation</u> subscale: *the app wasn’t very stimulating* (prompts - For example this could relate to how valuable, exciting, motivating or interesting you thought the app was or was not.)
- If scored low on <u>novelty</u> subscale: *the app wasn’t very novel* (prompts - For example, this could relate to how creative, cutting-edge, innovative, or inventive you thought the app was or was not.) *Could you describe your experience around this, or describe why you felt this was?*
- **Goals tracker feature:** I’d like to find out about your experience with the app’s specific features. Can you describe your experience of using the goals tracker feature? Did you like it or not like it? Why was this?

- If they didn’t like it: *What would you change / add?*
- **Mood tracker:** Can you describe your experience of using the mood tracker feature? Did you like it or not like it? Why was this?

- If they didn’t like it: *What would you change / add?*
- **Journal:** Can you describe your experience of using the journal feature? Did you like it or not like it? Why was this?

- If they didn’t like it: *What would you change / add?*
- **Immediate support:** Can you describe your experience of using the immediate support feature? Did you like it or not like it? Why was this?

- If they didn’t like it: *What would you change / add?*
- **Mascot:** Can you describe your experience of the mascot (dog)? Did you like it or not like it? Why was this?

- If they didn’t like it: *What would you change / add?*
- ***Expectations:*** *Before you started using the app, what were your expectations of what it should do?*

- *Was the app able to meet these expectations, or were any expectations not met?*
- *Did it have all the features you wanted or was anything missing?*
- *Did you use the app as much as you intended or not?*
- **General feedback:** Would you change anything about the app or not?
4. Integration of therapy and the app

- **Opening question:** We’re getting towards the end of the interview now. I’d like to talk a bit about how you found the therapy combined with the app. Can you describe a time when you worked together with the therapist on the app, e.g. discussing how to use the app?

- *What did the app add to the therapy, if anything? (Ask for examples)*
- ***Therapist support:*** *Can you describe your experience of the support the therapist provided around the app?* (prompts – Did you know you what you should do with the app and how it should work?)
5. Final reflection

- *“Is there anything else you’d like to discuss that we haven’t talked about?”*

***Stop recording***

### Appendix C: Participant demographics

**Table 1:**
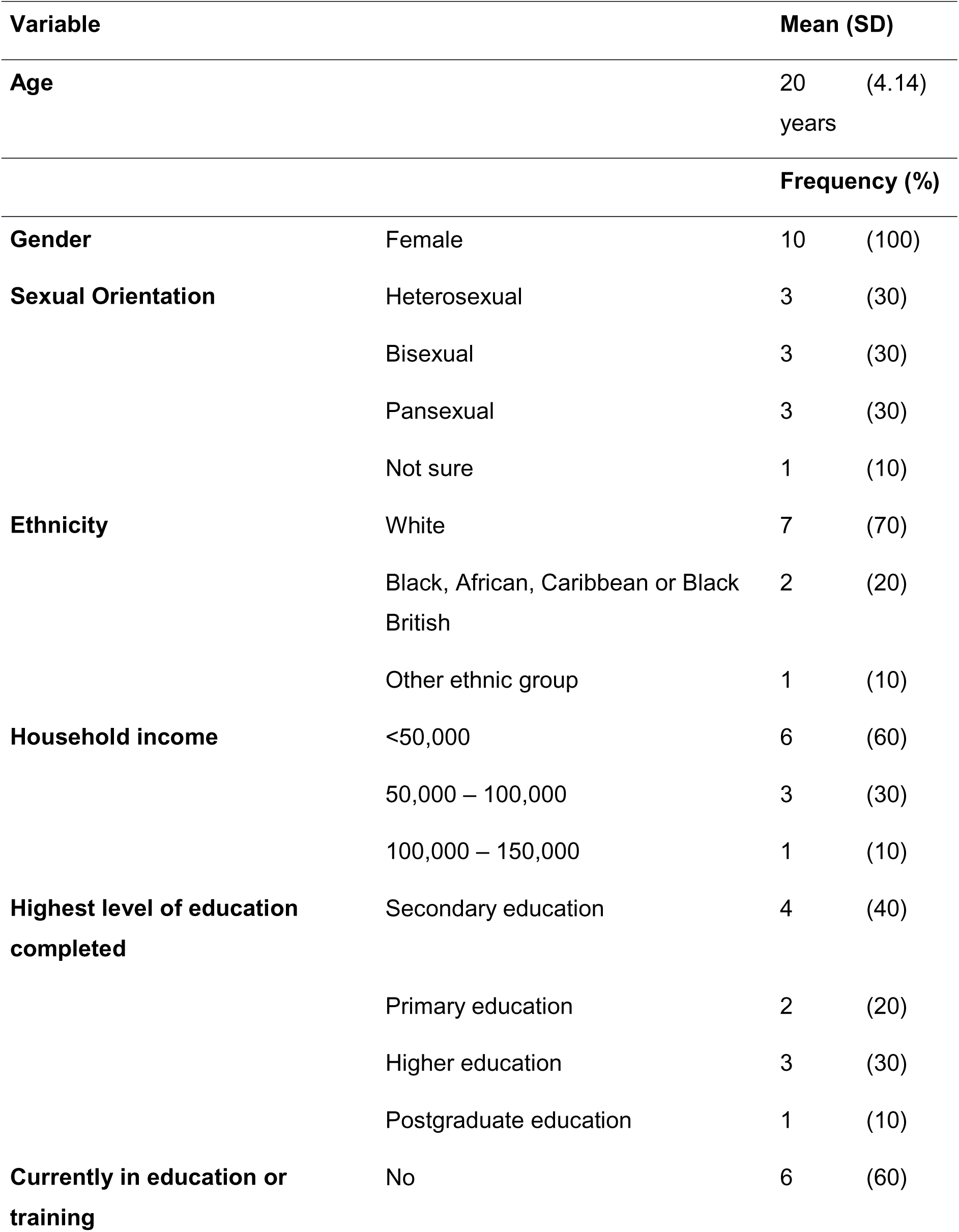
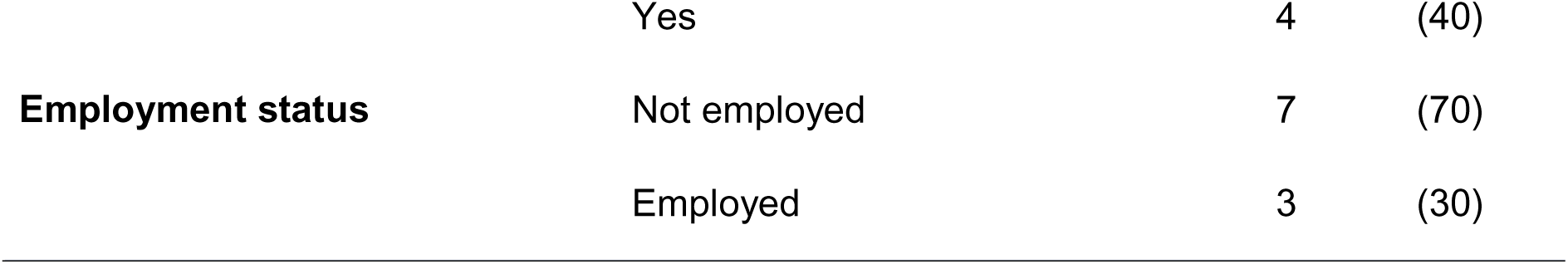
Participant demographics.

